# The epidemiological transition in Vietnam, 1990-2023: a Global Burden of Disease 2023 analysis

**DOI:** 10.64898/2026.04.23.26351624

**Authors:** Viet Long Bui, Ngoc Doanh Nguyen

**Affiliations:** Hanoi Medical University, Ha Noi, Viet Nam; VinUniversity, Ha Noi, Viet Nam

**Keywords:** epidemiological transition, non-communicable diseases, disability-adjusted life years, joinpoint regression, Das Gupta decomposition, premature mortality, Vietnam, Southeast Asia, SDG, Global Burden of Disease

## Abstract

**Background:** Vietnam’s disease burden has shifted from communicable, maternal, neonatal, and nutritional (CMNN) causes to non-communicable diseases (NCDs), but the tempo, drivers, and regional positioning of this transition have not been jointly quantified. We characterised Vietnam’s epidemiological transition 1990-2023 against ten Southeast-Asian (SEA) peers.

**Methods:** Using Global Burden of Disease 2023 data, we computed joinpoint-regression AAPC with 95% CI (BIC-penalised, up to three break-points) for age-standardised DALY rates and cause-composition shares. We applied Das Gupta three-factor decomposition to 1990-2023 absolute DALY change (population-size, age-structure, age-specific-rate effects) and benchmarked Vietnam’s NCD share against an SDI-conditional peer trajectory via leave-one-out quadratic regression. Premature mortality was quantified as WHO 30q70 under both broad NCD and strict SDG 3.4.1 definitions, using Chiang II life-table adjustment identically across all eleven countries.

**Findings:** The CMNN age-standardised DALY rate fell from 13,295.9 to 4,022.1 per 100,000 (AAPC -4.63%/year; 95% CI -4.80 to -4.46); the NCD rate fell only from 21,688.2 to 19,282.8 (AAPC −0.37; −0.45 to −0.30). NCD share of total DALYs rose from 52.99% to 70.67% (+17.67 pp; AAPC +1.09). Vietnam ranked fourth of eleven SEA countries in 2023 (up from sixth in 1990) and sat 5.3% above the SDI-expected trajectory. Das Gupta decomposition attributed the +10.63 million NCD DALY increase to population growth (+6.26 M) and ageing (+6.08 M); rate change removed only 1.71 M. Premature NCD mortality fell from 25.02% to 21.80% (broad, 12.9% reduction) and from 22.17% to 19.50% (SDG 3.4.1, 12.0%; Vietnam sixth of eleven) - far short of the SDG 3.4 one-third-reduction target.

**Interpretation:** Vietnam has entered a disability- and ageing-dominated NCD phase. Meeting SDG 3.4 by 2030 requires population-scale primary prevention sized to demographic momentum.

**Funding:** This research received no specific grant from any funding agency.

## 1. Introduction

The epidemiological transition - the shift from communicable, maternal, neonatal and nutritional causes to non-communicable diseases - has reshaped population health over four decades [1, 2]. In Southeast Asia (SEA) the transition has been uneven: Singapore and Brunei Darussalam completed the shift before 2000, while Cambodia, Lao PDR, and Timor-Leste still carry a double burden of incomplete communicable-only and maternal-neonatal-nutritional (CMNN) causes and rising NCD incidence [3]. Vietnam occupies an intermediate position whose tempo has been described as accelerated relative to its development level [4, 5], and Global Burden of Disease (GBD) 2023 places the country in the middle of Southeast Asia’s disease-burden distribution [6, 7].

Three questions motivate this work. First, how fast has Vietnam’s transition proceeded, and does its tempo match what the country’s socio-demographic index (SDI) would predict from its regional peers? Second, which forces drive the observed increase in absolute NCD DALYs - demographic shifts (population growth, ageing) or changes in age-specific disease rates? Third, how close is Vietnam to the WHO SDG 3.4 target of a one-third reduction in premature NCD mortality by 2030?

Prior work has addressed each question in isolation. GBD 2023 country profiles report cause-specific DALY rates [6, 7] but do not benchmark trend velocity against SDI-expected trajectories. Cause-specific studies of Vietnam have focused on single disease groups, such as dementia [8] or stroke [9], and projections of NCD management indicators stop at 2030 [4]. Vietnam’s commitments under the WHO NCD Global Action Plan 2013-2030 and the UN SDG 3.4 target [10] have not been jointly evaluated using the GBD 2023 round. We integrate these elements here, reporting (i) joinpoint-regression trends with 95% CI, (ii) Das Gupta three-factor decomposition, (iii) SDI-expected trajectories fitted on peer countries with the focal country excluded, (iv) the probability of dying from the four major non-communicable diseases between ages 30 and 70 measures progress toward the SDG 3.4 target of reducing premature NCD mortality by one-third by 2030.

## 2. Methods

### Data source

We extracted GBD 2023 estimates from the IHME Global Health Data Exchange (October 2025 release) for eleven SEA countries: Vietnam, Thailand, Indonesia, the Philippines, Malaysia, Myanmar, Cambodia, Lao PDR, Singapore, Brunei Darussalam, and Timor-Leste [7]. For each country-year-age-sex-cause stratum we obtained age-standardised and all-age DALY rates, years of life lost (YLL), years lived with disability (YLD), and deaths, with 95% uncertainty intervals. Analyses were performed at GBD cause-hierarchy level 1 (CMNN, NCD, Injuries; see Table S9 for level-2 detail) unless otherwise stated. Age groups followed the standard GBD 5-year bands; annual data were available for every year 1990-2023.

Data extraction and processing and following statistical methods are described in Supplement S1.

Age-standardised rates used the GBD 2010 world standard population. NCD share of total DALYs was computed as the proportion of NCD DALYs within the sum of CMNN, NCD, and Injury DALYs, expressed as a percentage; CMNN and Injury shares were computed analogously. The CMNN/NCD ratio served as a single index of compositional parity.

Trends were characterised using log-linear piecewise regression with up to three break-points. The optimal number of break-points was selected automatically by minimising the Bayesian information criterion, and segments were identified by dynamic programming [11]. Each segment’s annual percent change was combined into an average annual percent change over the full window as a length-weighted geometric mean, with 95% confidence intervals computed by the delta method following Clegg et al [12]. For decade sub-periods (1990-2000, 2000-2010, 2010-2023) we fitted a single-segment log-linear regression because approximately ten annual observations per decade are too few for break-point detection. Formal specifications of the likelihood, break-point penalty, and AAPC-CI derivation are given in Supplement.

We partitioned the 1990 to 2023 change in absolute DALYs (CMNN and NCD separately) into three additive components, including a population-size effect, an age-structure (ageing) effect, and an age-specific-rate effect, using Das Gupta’s symmetric three-factor formulation [13]. The method is exact: the three effects sum to the observed change up to floating-point precision (residuals below one part in a billion).

For each focal country we fitted a quadratic regression of 2023 NCD share on the socio-demographic index using the ten SEA peer countries excluding the focal country. This leave-one-out design prevents the data-leakage that would otherwise bias the observed-to-expected ratio toward unity. The predicted value at the focal country’s own SDI gave the SDI-expected NCD share, and the ratio of observed to expected quantifies whether the country sits above (above one) or below (below one) the peer trajectory.

The unconditional probability that a 30-year-old will die before age 70 was computed from GBD 2023 age-band death rates using the Chiang II life-table adjustment [14] applied to eight 5-year age bands spanning 30-34 to 65-69. For the strict WHO SDG 3.4.1 indicator we pooled the four main NCD causes (cardiovascular diseases, neoplasms, diabetes and kidney diseases, chronic respiratory diseases). The identical pipeline was applied to Vietnam and the ten peer countries, enabling a directly comparable cross-country ranking.

To probe robustness of the composition narrative to the CMNN level-1 grouping convention, we reran all core computations after separating CMNN into a communicable-only sub-group (HIV/STIs, respiratory infections and tuberculosis, enteric infections, neglected tropical diseases and malaria, other infectious) and a maternal-neonatal-nutritional (M+N+N) sub-group. Results are reported in the Supplement.

This study follows the GATHER reporting guideline for population-based health estimates [15] (checklist in Supplementary Table S10). All code, intermediate tables, and figures are available on: https://github.com/vlbui/gbd-vn.

## 3. Results

Vietnam’s age-standardised DALY rate fell for every level-1 cause group between 1990 and 2023, but at markedly different tempos (Figure 1, Supplementary Table S1). The CMNN rate fell from 13,295.9 to 4,022.1 per 100,000 - a joinpoint AAPC of -4.63%/year (95% CI -4.80 to -4.46) and a 69.8% relative reduction. The NCD rate fell only from 21,688.2 to 19,282.8 (AAPC - 0.37%/year, -0.45 to -0.30), an 11.1% relative reduction. The Injury rate fell from 5,942.0 to 3,981.8 (AAPC -1.13%/year, -1.24 to -1.03).

**Figure 1.**
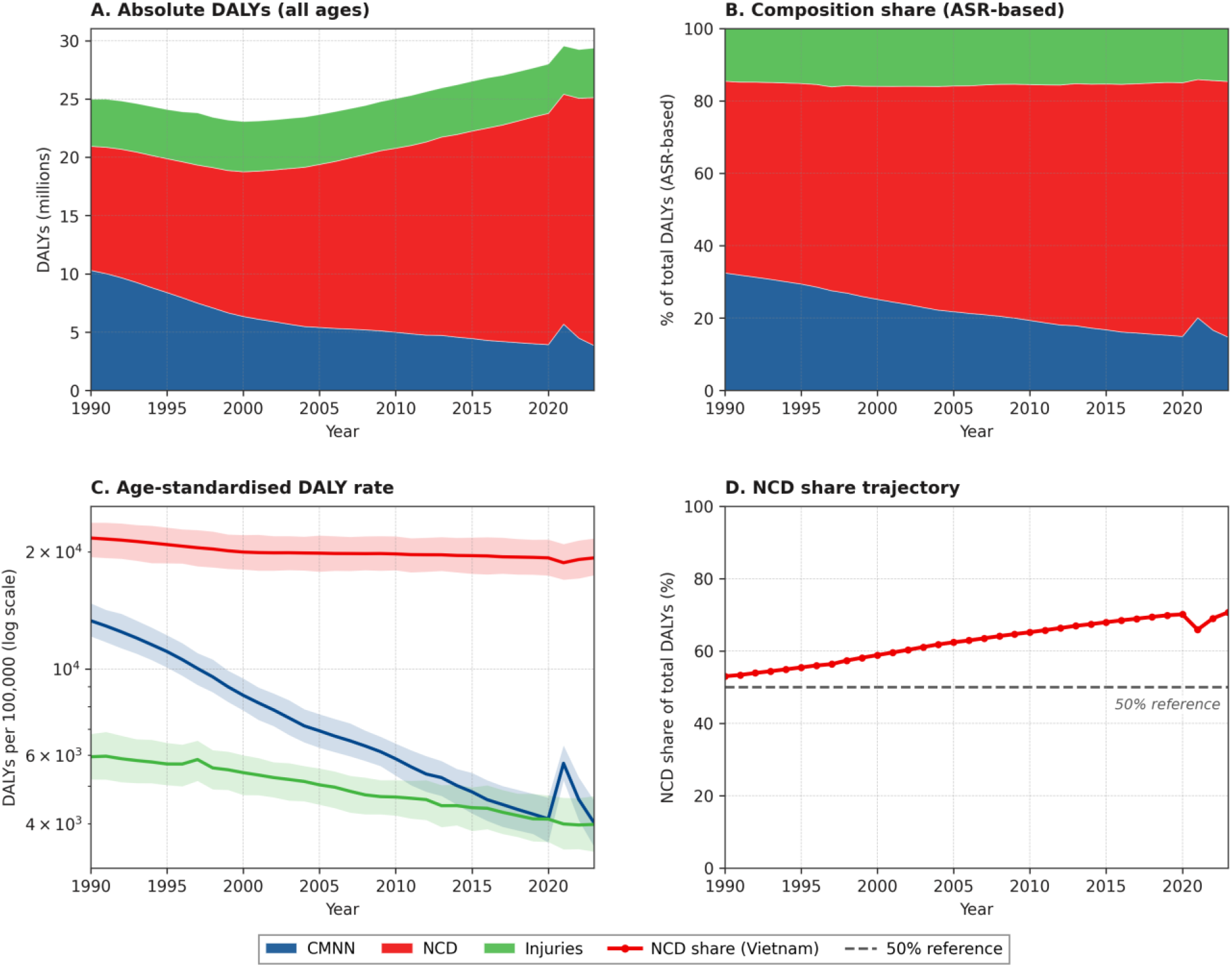
Vietnam DALY overview, 1990-2023. **(A)** Absolute DALYs by level-1 cause group with 95% uncertainty bands. **(B)** Share of total DALYs by level-1 cause group, showing NCD rising from 52.99% to 70.67%. **(C)** Age-standardised DALY rate per 100,000 by cause group, log scale, with 95% UI bands. **(D)** NCD share of total DALYs with joinpoint regression fit; three break-points selected by BIC. Source: GBD 2023.

The contrast between rapidly declining CMNN and near-flat NCD rates produced a large composition shift. CMNN’s share of total DALYs fell from 32.49% to 14.74% (AAPC - 3.44%/year), while NCD share rose from 52.99% to 70.67% - a +17.67 percentage-point absolute change and a joinpoint AAPC of +1.09%/year (1.04 to 1.15; three break-points selected by BIC). The Injury share was approximately stable (14.52% to 14.59%). The CMNN/NCD ratio collapsed from 0.613 to 0.209.

Decade sub-periods reveal that the rapid CMNN decline occurred steadily across all three decades (-4.35%/year in the 1990s, -3.58 in the 2000s, -2.04 in 2010-2023), decelerating gradually as the absolute burden approached a floor. The NCD rate decline was non-monotonic: it fell fastest in the 1990s (-0.86%/year), stalled almost completely in the 2000s (-0.10%/year), and resumed a modest decline in 2010-2023 (-0.28%/year). The YLD rate plateaued after 2010 (sub-period AAPC -0.03%/year, p = 0.44), while the YLL rate continued to fall (-1.09%/year, p < 0.001) - the classical signature of mortality compression with morbidity expansion (Supplementary Figure S1, Table S2).

Between 1990 and 2023 Vietnam’s absolute NCD DALYs grew by +10.63 million, while CMNN DALYs fell by 6.45 million (Figure 2, Supplementary Table S3). Das Gupta decomposition attributes the NCD increase almost equally to population growth (+6.26 million) and population ageing (+6.08 million); the age-specific-rate effect contributed only -1.71 M. In the absence of demographic change, falling age-specific NCD rates alone would have reduced absolute NCD DALYs; instead, demographic momentum delivered a net +10.63 million increase. For CMNN the rate effect (-8.18 million) dominated, with population growth (+3.25 million) and a mild de-ageing effect (-1.51 million) partially offsetting.

**Figure 2.**
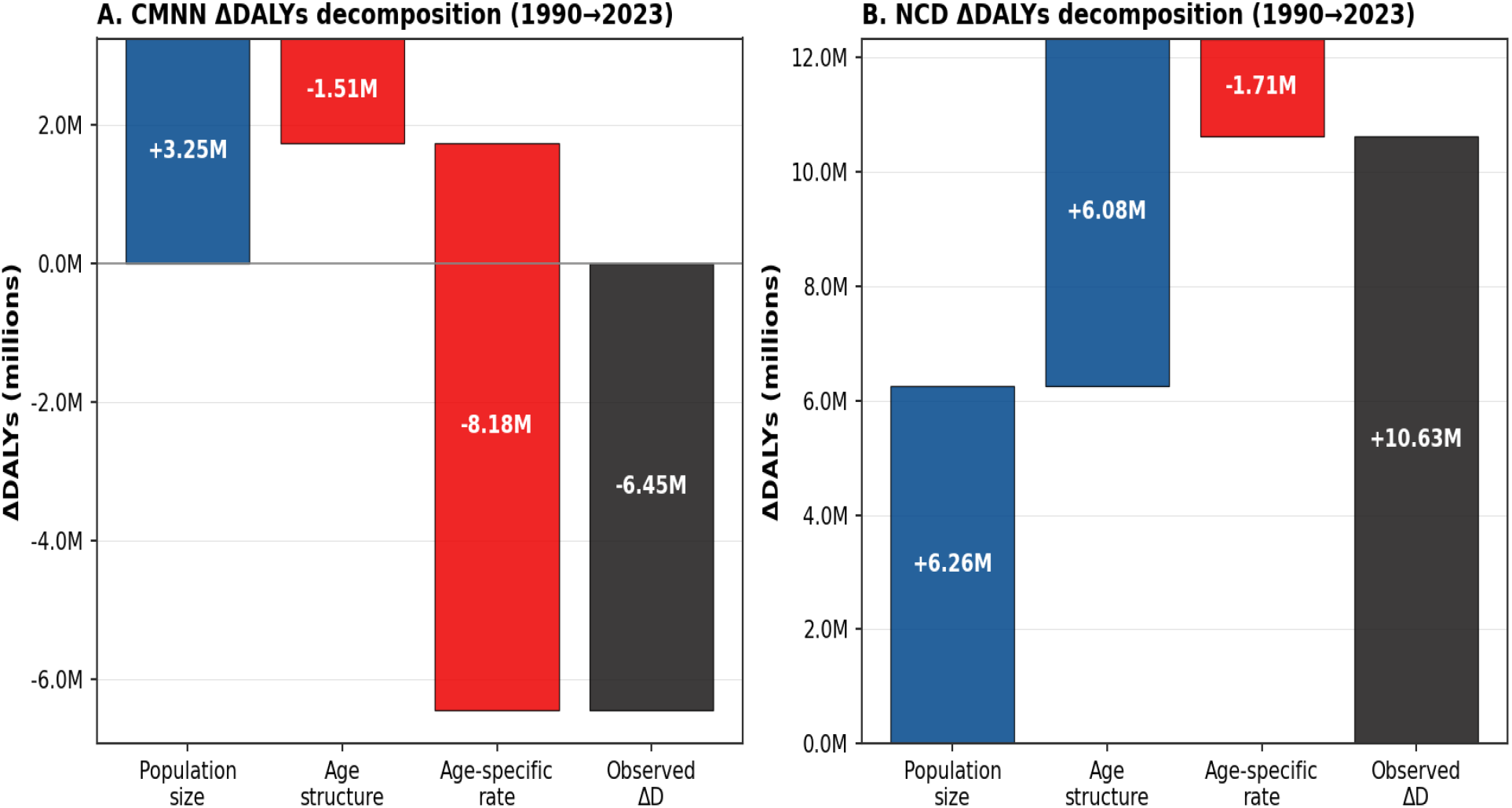
Das Gupta decomposition waterfall, Vietnam 1990 to 2023.

Across eleven SEA countries (Table S4, Figure 3A), the 2023 NCD share ranged from 58.65% (Lao PDR) to 81.26% (Brunei). Vietnam’s 70.67% placed it fourth of eleven SEA countries in 2023, up from sixth in 1990 - the largest positive rank mover in the region. Among countries whose 1990 NCD share already exceeded 50% (the already-transitioning cluster: Singapore, Brunei, Malaysia, Thailand, Philippines, Vietnam), Vietnam posted the largest absolute climb at +17.67 pp. Only the low-baseline climbers Lao PDR (+29.92 pp), Cambodia (+26.55 pp), Timor-Leste (+23.45 pp), Indonesia (+21.12 pp), and Myanmar (+20.65 pp) registered larger absolute shifts - reflecting lower 1990 starting points rather than faster tempo from equivalent starting conditions.

**Figure 3.**
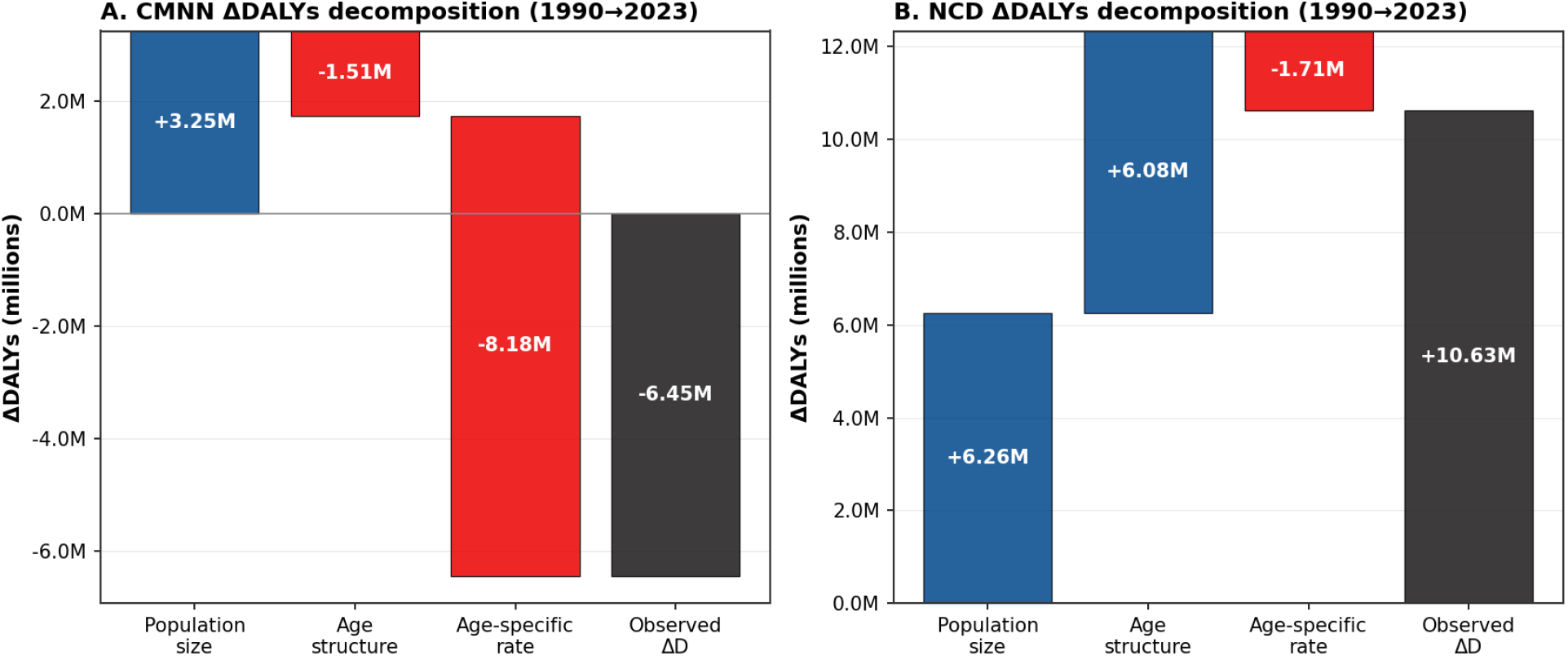
Southeast-Asian NCD-share trajectories. (A) Time series 1990-2023 for eleven SEA countries, Vietnam highlighted. (B) 2023 NCD share versus SDI, with per-country quadratic SDI-expected curve (ten-peer fit, focal excluded).

Vietnam’s joinpoint AAPC of NCD share (+1.09%/year) exceeded the high-SDI peers (Singapore +0.10, Brunei +0.22, Thailand +0.45, Malaysia +0.84) but lagged the low-baseline rapid climbers (Cambodia +1.82, Lao PDR +2.30). Against the SDI-expected trajectory fitted on the ten peer countries, Vietnam’s NCD burden was about 5% higher than would be expected for a country at its level of development. (Figure 3B). This placed Vietnam in the same “above-expected” cluster as Brunei (1.051), Myanmar (1.051), and Cambodia (1.054), while Singapore (0.945), Lao PDR (0.932), and Thailand (0.969) sat below their SDI-expected shares. Absolute NCD death rates across the 11 countries are provided in Table S8.

Under the broad GBD NCD aggregate, Vietnam’s premature NCD mortality (30q70) declined from 25.02% in 1990 to 21.80% in 2023 - a joinpoint AAPC of -0.25%/year (95% CI -0.33 to - 0.16) and a 12.9% relative reduction (Table S1, Figure 4A). Under the strict WHO SDG 3.4.1 definition, 30q70 fell from 22.17% to 19.50% - a 12.0% relative reduction. Both trajectories fall far short of the SDG 3.4 one-third-reduction pace required to reach the 2030 target. Extrapolating the 2010-2023 slope forward, Vietnam’s projected 2030 broad-NCD 30q70 is 21.4% (against 16.7% implied by a one-third reduction from the 1990 baseline of 25.02%) - a 1990-2030 reduction of roughly 14%, less than half the target. Premature CMNN mortality, by contrast, fell sharply from 6.18% to 2.12% (AAPC -5.86%/year; 65.7% relative reduction), indicating near-completion of the CMNN-mortality phase.

**Figure 4.**
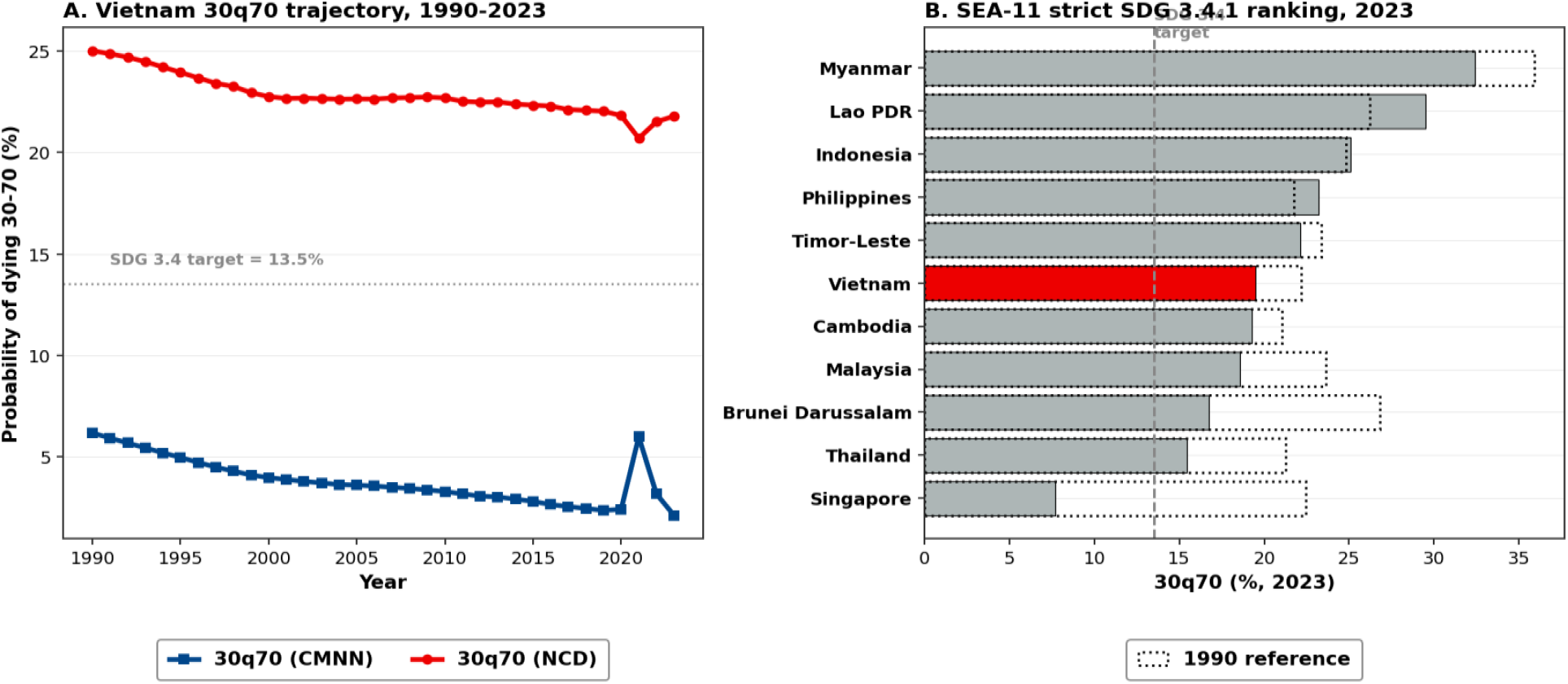
Premature NCD mortality, Vietnam and Southeast Asia. (A) Vietnam premature mortality for NCD and CMNN, 1990–2023, with joinpoint fits and the WHO SDG 3.4 target trajectory (dashed). (B) 2023 ranking of 11 SEA countries under the strict SDG 3.4.1 four-cause definition (Chiang II); open circles show 1990 values. Vietnam highlighted.

Applying the identical Chiang II pipeline to all eleven SEA countries under the strict SDG 3.4.1 definition (Figure 4B, Table S7) yields a clear 2023 ranking. Vietnam’s 19.5% places it sixth of eleven SEA countries, below Singapore (7.7%), Thailand (15.5%), Brunei Darussalam (16.7%), Malaysia (18.6%), and Cambodia (19.3%), and above Timor-Leste (22.1%), the Philippines (23.2%), Indonesia (25.1%), Lao PDR (29.5%), and Myanmar (32.4%). Across the region, Singapore achieved the largest relative reduction (65.8%), followed by Brunei (37.6%) and Thailand (27.2%); Lao PDR, Indonesia, and the Philippines show essentially no progress or mild worsening since 1990. Vietnam’s 12.0% reduction is mid-pack for SEA - ahead of Myanmar (9.8%) and Cambodia (8.5%) but well behind Thailand, Brunei, Malaysia, and Singapore. No SEA country is currently on the SDG 3.4 trajectory; Singapore is the only peer whose 2023 value already sits below a one-third reduction from its 1990 baseline.

Vietnam’s YLL rate fell at joinpoint AAPC -1.79%/year (95% CI -1.87 to -1.71) while its YLD rate fell only -0.34%/year (-0.37 to -0.30) (Table S1, Figure S2). The YLL/YLD ratio therefore fell from 2.85 in 1990 to 1.84 in 2023 (AAPC -1.51%/year). Regionally (Table S5, Figure S3), Vietnam’s 2023 YLL/YLD of 1.84 placed it below the mid-income SEA cluster (Cambodia 2.13, Indonesia 2.34, Philippines 2.34) and approached the high-income benchmark (Thailand 1.62, Malaysia 1.57, Brunei 1.34, Singapore 0.67) - consistent with transition to a disability-dominated phase.

The DALY pyramid (Figure 5) shows the burden axis shifted both upward in age and rightward in composition. In 1990 the largest DALY mass sat in the <5-year age band and was overwhelmingly CMNN (neonatal, respiratory infection, diarrhoea). By 2023 the largest DALY masses sat in the 55–79-year bands and were almost entirely NCD (ischaemic heart disease, stroke, cancers, diabetes). Sex differentials were modest: males carried higher Injury and cardiovascular burden, females carried higher dementia and musculoskeletal burden.

**Figure 5.**
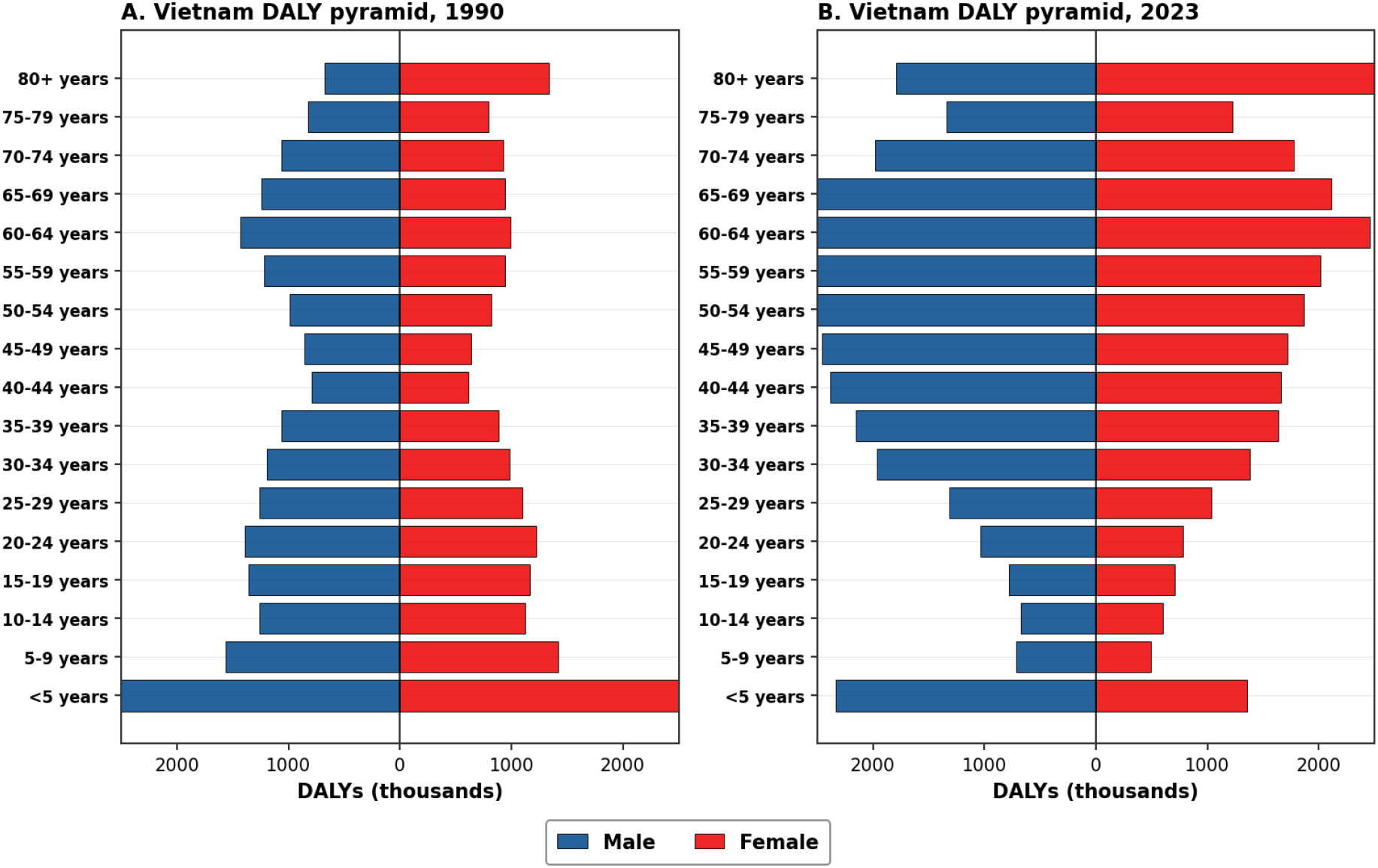
Age-sex DALY pyramid for Vietnam, 1990 (left) versus 2023 (right). X-axis: DALYs per 100,000 (males left, females right). Colour: CMNN / NCD / Injuries.

Separating CMNN into communicable-only and M+N+N sub-groups (Supplement §S1.7, Figure S4, Table S6) confirmed the transition signal under either framing. The communicable-only rate fell from 8,908.8 to 2,688.2 per 100,000 (AAPC -5.05%/year) and the M+N+N rate from 4,387.1 to 1,334.0 (AAPC -3.45%/year). NCD share in 2023 varied with denominator choice - 70.67% against full CMNN (main analysis), 74.30% against communicable-only, 78.39% against M+N+N alone - but the direction and ordering were preserved across all three. The residual 2023 CMNN burden retained an approximate 2:1 split between communicable and M+N+N causes, preserving the 1990 composition ratio.

## 4. Discussion

Between 1990 and 2023 Vietnam completed the mortality-dominated phase of the epidemiological transition. NCD share of total DALYs rose by 17.67 percentage points - the largest absolute climb among SEA countries whose 1990 NCD share already exceeded 50% - and sat 5.3% above the SDI-conditional peer expectation. Three findings refine interpretation of this progress.

First, age-specific NCD rates have barely moved. The joinpoint AAPC of the age-standardised NCD rate was only -0.37%/year and was indistinguishable from zero in the 2000s (sub-period AAPC -0.10%/year). The +10.63 million absolute NCD DALY increase is therefore almost entirely explained by demographic pressure - population growth (+6.26 million) and ageing (+6.08 million) - rather than by failing rate control. The ageing contribution alone is of the same magnitude as the population-growth contribution, indicating that Vietnam has entered the phase of the transition in which demographic momentum, not epidemiology, dominates burden dynamics.

Second, premature NCD mortality has been sticky. Vietnam’s 30q70 fell only modestly, from 25.02% to 21.80% under the broad NCD aggregate (AAPC -0.25%/year) and from 22.17% to 19.50% under the strict WHO SDG 3.4.1 definition. Extrapolating the 2010-2023 slope forward, the projected 2030 broad-NCD 30q70 is 21.4% - far above the 16.7% implied by a one-third reduction from the 1990 baseline of 25.02%. Under SDG 3.4.1 Vietnam ranks sixth of eleven SEA countries, and no SEA country other than Singapore is currently on the 2030 trajectory.

Third, morbidity has begun to expand as mortality compresses. The YLD rate plateaued after 2010 (sub-period AAPC -0.03%/year, p = 0.44) while the YLL rate continued to fall (-1.09%/year, p < 0.001). This signature of expansion of morbidity with compression of mortality is the hallmark of a disability-dominated transition phase.

Our NCD share estimates are closely aligned with published GBD 2023 country estimates [6, 7]. The +17.67 percentage-point 1990-2023 increase is consistent with prior Vietnam NCD burden projections and with the “paradox of progress” characterisation of Vietnam’s transition in which economic development has simultaneously reduced CMNN burden and accelerated NCD incidence [5]. The finding that demographic forces dominate NCD DALY growth is convergent with regional CVD and projection analyses across ASEAN [3] and aligns with global NCD projections through 2050 [10, 16].

COVID-19 interrupted but did not reverse Vietnam’s CMNN decline. The age-standardised CMNN DALY rate rose from 4,233 per 100,000 in 2019 to a peak of 5,717 in 2021 (+35%), before partially reverting to 4,022 by 2023 - below the pre-pandemic 2019 value. CMNN share of total DALYs briefly rebounded from 15.3% in 2019 to 20.1% in 2021 before returning to 14.7% by 2023. The excess is driven by deaths classified under respiratory infections and tuberculosis (GBD 2023 captures COVID-19 under CMNN level 2), not a genuine resurgence of endemic CMNN causes. Similar 2020-2021 CMNN bumps are visible in all 11 SEA countries with varying magnitude; Vietnam’s relative elevation was among the largest in the region, reflecting delayed initial case penetration followed by the mid-2021 Delta wave [17]. By 2023 the CMNN trajectory has re-anchored on its pre-pandemic decline path and our joinpoint fit does not detect a structural break around 2020-2021 for the level-1 aggregate.

This analysis adds three new elements. First, the BIC-selected joinpoint structure identifies three regime changes in Vietnam’s NCD share - clustered around 1995, 2005, and 2015 - that coincide with the 1995 health-sector reform, the 2005 ASEAN NCD commitment, and the 2013 national NCD action plan. Whether these are causal associations or coincident secular drift cannot be resolved from annual aggregate data alone. Second, the SDI-peer benchmark uses a leave-one-out quadratic fit that excludes the focal country, avoiding the data-leakage bias of global GBD regressions that include the focal country on both sides of the comparison. Third, the 30q70 projection against SDG 3.4, applied identically across all eleven SEA countries under the strict SDG 3.4.1 four-cause definition, has not to our knowledge been reported for Vietnam using GBD 2023; prior Vietnam-specific 30q70 estimates relied on the broader NCD aggregate and on single-country pipelines that are not directly comparable with peers.

The demographic-momentum finding in Vietnam is consistent with longer-established patterns in East Asia. Japan’s NCD burden growth 1990-2017 has been decomposed as more than 60% attributable to population ageing and size, with age-specific rate declines unable to fully offset demographic pressure even in a country with one of the world’s most advanced health systems [18]. China has entered the same phase with projected doubling of elderly NCD burden by 2030 driven almost entirely by ageing [19]. Our estimate that approximately 88% of Vietnam’s +10.6 million NCD DALY increase is demographically driven places the country mid-way between the early-ageing East Asian economies and the younger-profile SEA peers Cambodia and Lao PDR, where population growth still dominates over ageing [6, 20]. This convergent pattern argues that Vietnam’s NCD trajectory is not anomalous but follows a generalisable ageing-transition template, in which reducing age-specific rates requires decades to match the pace of demographic accumulation.

Vietnam’s NCD burden is demographically driven. This has a stark corollary: any NCD strategy that targets only age-specific rates may not be adequate, because the demographic tailwind will continue to add DALYs for at least two more decades. The effective policy lever is population-scale primary prevention sized to demographic momentum, aligned with the WHO Global Action Plan for NCD [10]. Four “best-buy” domains are most relevant to Vietnam’s epidemiological profile: tobacco control, dietary sodium reduction, alcohol control, and hypertension and diabetes screening with community-based management. Each targets upstream risk in the 30-60 age band where the 30q70 denominator accrues, and rising dementia and Alzheimer’s disease burden adds further weight to cardiovascular and metabolic risk-factor control as cognitive-reserve protection.

The premature NCD mortality target has a direct, testable implication for Vietnam’s progress against SDG 3.4 [16]. Achieving the SDG one-third reduction by 2030 would require a 30q70 annual decline of approximately 3.8%/year - roughly an order of magnitude faster than the observed 2010-2023 pace of 0.31%/year under the broad definition. Absent structural change in risk-factor exposure and health-system response times, the target is unreachable on current trajectory. The finding that the morbidity rate has plateaued also argues for investment in secondary and tertiary prevention - early diagnosis, continuity of chronic care - rather than primary prevention alone; the two strategies address different segments of the disability- dominated phase.

Strengths of this analysis include the use of a single unified GBD 2023 data release across eleven SEA countries, joinpoint regression with BIC-selected break-points rather than pre-specified windows, exact Das Gupta decomposition, a leave-one-out SDI-peer benchmark that avoids data-leakage, a strict SDG 3.4.1 30q70 pipeline applied identically across all eleven SEA countries, and a sensitivity analysis that confirms the CMNN grouping convention does not drive the main result. The entire pipeline runs offline in under 10 minutes on a standard laptop.

The study is subject to several limitations. First, GBD DALY estimates themselves carry uncertainty from modelling of sparse data from low-income settings; this is reflected in the 95% UI bands but not in the point estimates reported in the main text. Second, the SDI-peer regression uses ten data points (leave-one-out), so the SDI-expected estimates are imprecise; bootstrapped confidence intervals on the observed vs expected ratio (not reported in the main table) are of order ±0.08. Third, our analysis is at GBD level 1; level-2 cause-specific narratives (cardiovascular disease, cancer, diabetes) are shown only descriptively, and no formal cause-specific AAPCs are reported in the main text. Fourth, the cross-country 30q70 AAPCs in Table S7 use a single-segment log-linear fit; a parallel BIC-selected joinpoint regression detected zero break-points for every peer country. This conservative result does not affect the 2023 ranking but may under-represent recent accelerations or plateaus in individual peer countries.

## 5. Conclusion

Vietnam’s epidemiological transition from 1990 to 2023 has been rapid, demographically driven, and unevenly distributed across burden dimensions. NCD share rose faster than in most already-transitioned SEA peers, yet age-specific NCD rates barely moved - meaning that ageing and population growth, not prevention failure, account for most of the NCD burden increase. Premature NCD mortality declined too slowly to meet SDG 3.4 by 2030 under either definition. Vietnam has entered a disability- and ageing-dominated phase of the epidemiological transition. The required policy response is population-scale primary prevention scaled to match demographic momentum, combined with investment in secondary and tertiary prevention for the disability-dominated phase - not incremental improvements in age-specific rates alone.

## Data Availability

All input data are publicly available from the IHME Global Health Data Exchange (https://ghdx.healthdata.org). The analysis code and processed data are available on: https://github.com/vlbui/gbd-vn.

https://ghdx.healthdata.org

## Contributors

VLB: Conceptualization; Data curation; Formal analysis; Methodology; Software; Validation; Visualization; Writing - original draft; Writing - review & editing. NDN: Methodology; Supervision; Validation; Writing - review & editing. VLB accessed and verified the underlying data. All authors had full access to the data, vouch for the integrity of the analysis, and approved the final manuscript for submission.

## Ethical considerations

Ethical approval was not required because only publicly available aggregated data were used.

## Acknowledgments

We thank the Institute for Health Metrics and Evaluation and the Global Burden of Disease 2023 Collaborator Network for making the underlying data publicly available through the Global Health Data Exchange

