## Supplementary Material for "The epidemiological transition in Vietnam, 1990-2023: a Global Burden of Disease 2023 analysis"

### S1. Detailed methods

#### S1.1 Data source and extraction

We extracted Global Burden of Disease 2023 (GBD 2023) estimates from the IHME Global Health Data Exchange (GHDx), October 2025 release, through the GBD Results tool (https://vizhub.healthdata.org/gbd-results/). Eleven Southeast-Asian (SEA) countries were included: Vietnam (IHME location ID 20), Thailand (18), Indonesia (11), the Philippines (16), Malaysia (15), Myanmar (14), Cambodia (10), Lao PDR (12), Singapore (19), Brunei Darussalam (7), and Timor-Leste (369).

For each country-year-age-sex-cause stratum we obtained age-standardised and all-age DALY rates, years of life lost (YLL), years lived with disability (YLD), and deaths, together with 95% uncertainty intervals (2.5th and 97.5th percentiles). Cause-specific estimates originate from the Cause of Death Ensemble model (CODEm) and the DisMod-MR 2.1 Bayesian meta-regression framework. Primary analyses used GBD cause-hierarchy level 1 (CMNN, NCD, Injuries). Age groups followed GBD 5-year bands; annual data covered every year 1990-2023 for all analyses, and 1980-2023 for the 30q70 pipeline only.

#### S1.2 Age-standardisation and composition metrics

Age-standardised rates used the GBD 2010 world standard population. The NCD share of total DALYs was computed as:

$$\text{NCD share} = DALY_{NCD} / (DALY_{CMNN} + DALY_{NCD} + DALY_{Injury})$$

CMNN and Injury shares were computed analogously. The CMNN/NCD ratio served as a single index of compositional parity.

#### S1.3 Joinpoint regression and AAPC

Temporal trends were characterised using log-linear piecewise regression of the annual age-standardised rate (or share). For segment k:

$$\log y_t = \alpha_k + \beta_k \cdot t, \quad t \in \text{segment } k$$

Up to three break-points were allowed. The optimal number of break-points was selected by minimising the Bayesian information criterion (BIC):

$$\text{BIC} = n \cdot \log(\text{SSR} / n) + p \cdot \log(n)$$

where n is the number of annual observations, SSR is the residual sum of squares, and p counts the free parameters (2 × number of segments). Segments were identified by dynamic programming.

The annual percent change in segment k:

$$\text{APC}_k = 100 \cdot (\exp(\beta_k) - 1)$$

The average annual percent change (AAPC) over the full window is the length-weighted geometric mean of segment APCs:

$$\text{AAPC} = 100 \cdot [\ \textstyle\prod_k (1 + \text{APC}_k / 100)^{Lk\,/\,L} - 1\ ]$$

where $L_k$ is segment length in years and L is total window length.

95% confidence intervals were computed by the delta method following Clegg et al. 2009 (Stat Med).

$$95\%\ \text{CI} = \text{AAPC} \pm 1.96 \cdot \text{SE(AAPC)}, \quad \text{SE(AAPC)} \approx 100 \cdot \exp(\beta) \cdot \text{SE}(\beta)$$

For decade sub-periods we fitted single-segment log-linear regression.

#### S1.4 Das Gupta three-factor decomposition

Absolute DALY change from 1990 to 2023 was partitioned into three additive components using Das Gupta's symmetric formulation:

$$\Delta D = (\Delta P) \cdot \bar{s} \cdot \bar{r} \quad \text{[population-size effect]}$$

$$+ \bar{P} \cdot (\Delta s) \cdot \bar{r} \quad \text{[age-structure / ageing effect]}$$

$$+ \bar{P} \cdot \bar{s} \cdot (\Delta r) \quad \text{[age-specific rate effect]}$$

P is total population size, s is the age-structure vector, r is the vector of age-specific DALY rates; barred quantities are 1990/2023 averages. The method is exact: components sum to observed change up to $10^{-9}$ precision.

#### S1.5 SDI-benchmarked expectation (leave-one-out)

For each focal country c we fitted a quadratic regression of 2023 NCD share on SDI using the ten SEA peer countries, excluding the focal country:

$$\text{NCD share}_i = \beta_0 + \beta_1 \cdot \text{SDI}_i + \beta_2 \cdot \text{SDI}_i^2 + \varepsilon_i, \quad i \neq c$$

$$\text{obs / exp}_c = \text{NCD share}_c \,/\, \text{NCD share}_{\text{expected, c}}$$

#### S1.6 Premature mortality: Chiang II 30q70

30q70 was computed from GBD 2023 age-band death rates using the Chiang II abridged life-table adjustment (Eayres & Williams 2004):

$$q_a = (5 \cdot m_a) / (1 + 2.5 \cdot m_a)$$

$${}_{30}q_{70} = 1 - \prod_{a \in \{30, 35, ..., 65\}} (1 - q_a)$$

Strict WHO SDG 3.4.1 uses the 4 main NCDs pooled; broad NCD uses the full GBD level-1 NCD aggregate.

#### S1.7 CMNN sub-group sensitivity

CMNN was separated into communicable-only (HIV/STIs, respiratory infections and TB, enteric, NTDs/malaria, other infectious) and maternal-neonatal-nutritional (M+N+N). NCD shares recomputed against each sub-denominator.

#### S1.8 Reproducibility

Code (Python 3.10, NumPy, pandas, Plotly; joinpoint via dynamic programming / ruptures) runs end-to-end in under 10 minutes. Follows GATHER reporting guideline (Table S10). Only publicly available aggregated data used.

### S2. Additional results

#### S2.1 Supplementary figures

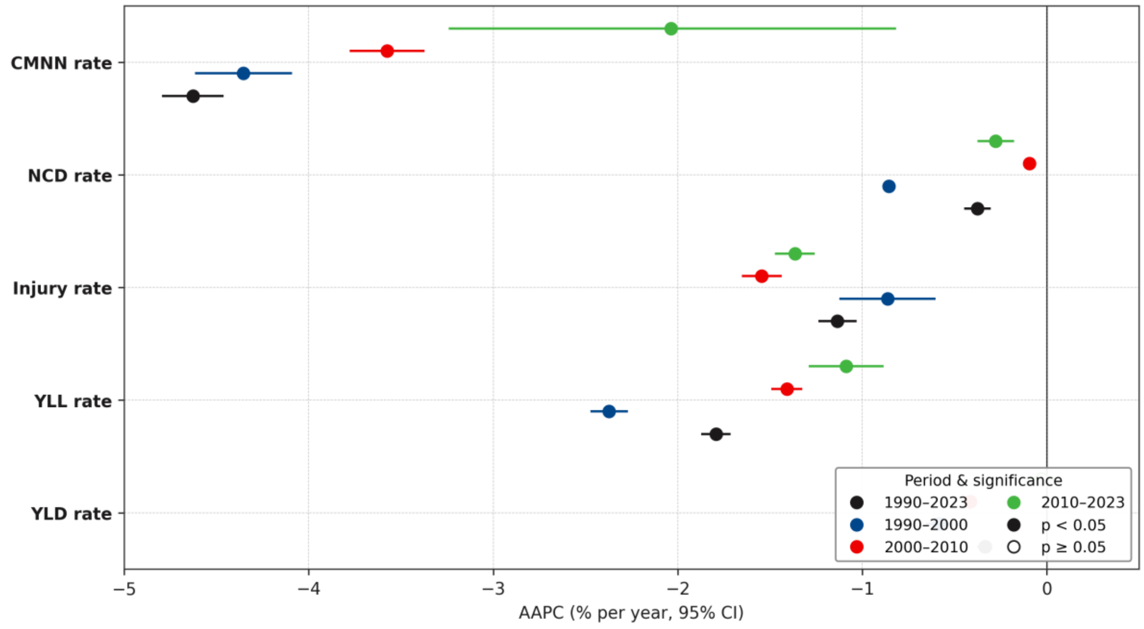

**Figure S1.** Period-specific AAPC forest plot for five burden metrics (age-standardised CMNN, NCD, Injury DALY rates; YLL rate; YLD rate), Vietnam 1990-2023. Dots = point estimates; bars = 95% CI; colour = time window (full 1990-2023 black, 1990s blue, 2000s red, 2010-2023 green). Open markers flag sub-periods with $p \geq 0.05$.

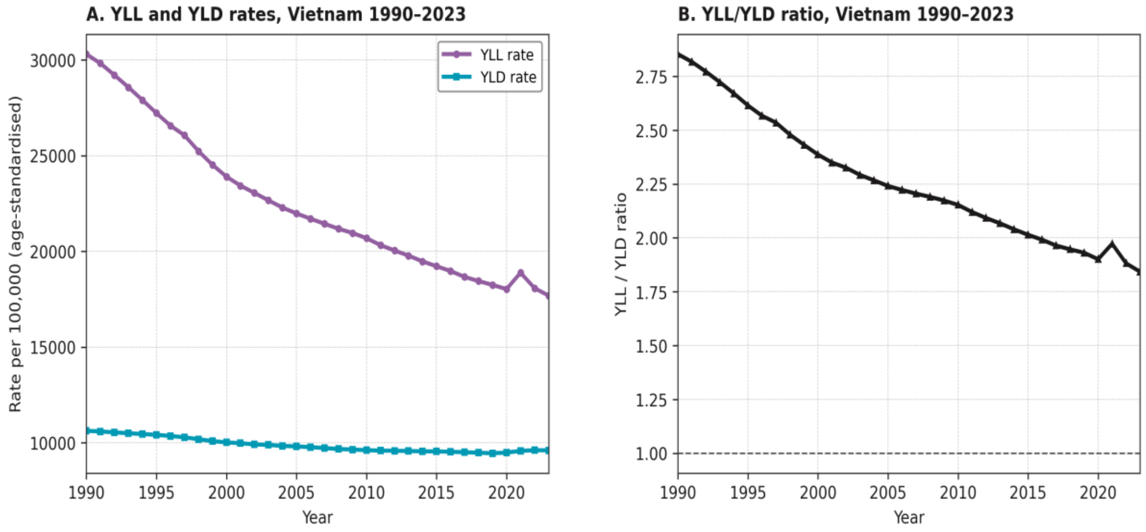

**Figure S2.** Vietnam YLL rate, YLD rate, and YLL/YLD ratio, 1990-2023. Joinpoint fits overlaid. YLD plateau after 2010 (AAPC -0.03%/year, $p = 0.44$) vs continued YLL fall (-1.09%/year, $p < 0.001$).

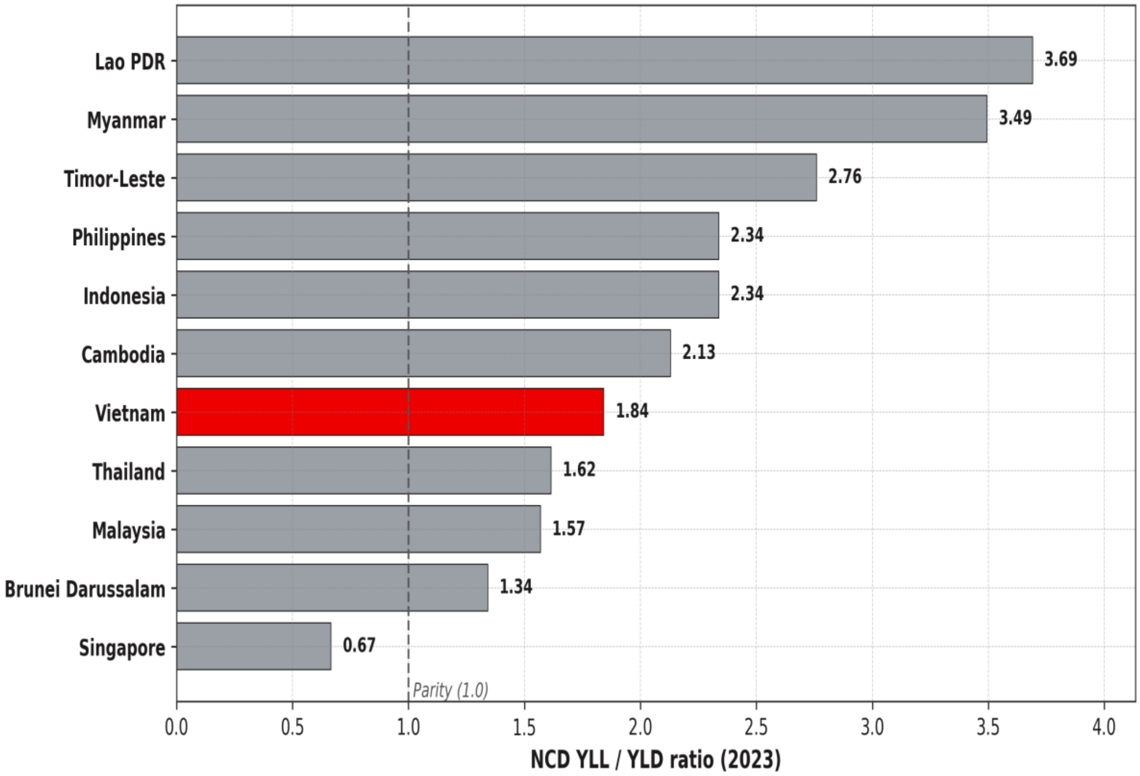

**Figure S3.** Southeast-Asian YLL/YLD ratio ranking, 2023. Vietnam's 1.84 sits between mid-income cluster (Cambodia 2.13, Indonesia/Philippines 2.34) and high-income benchmark (Thailand 1.62, Malaysia 1.57, Brunei 1.34, Singapore 0.67).

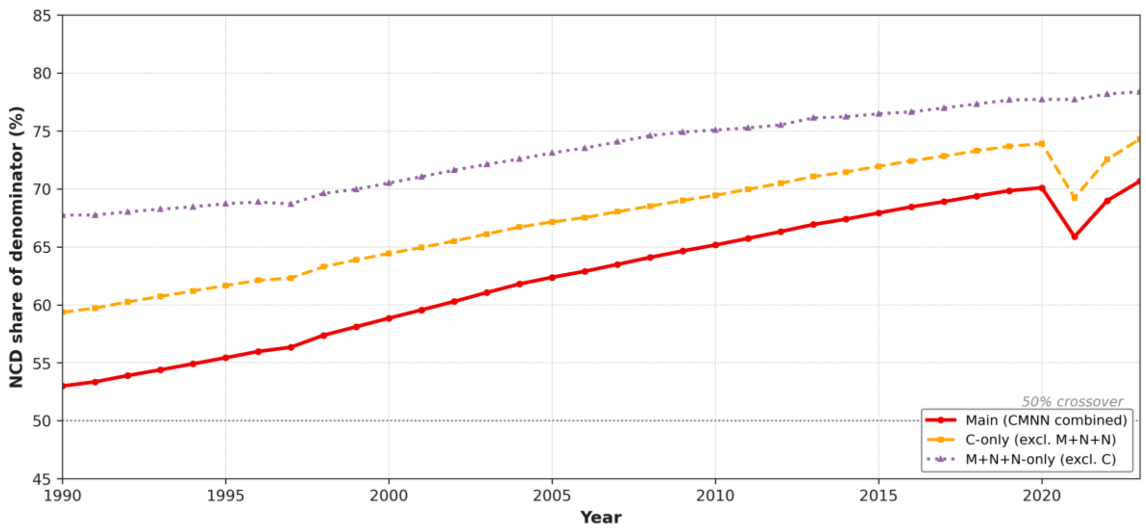

**Figure S4.** CMNN grouping sensitivity: full CMNN, communicable-only, and M+N+N sub-groups, Vietnam 1990-2023. Transition signal preserved under every sub-denominator choice.

#### S2.2 Supplementary tables

**Table S1.** Vietnam key-metric summary, 1990-2023. Age-standardised DALY rates (per 100,000) by level-1 cause group; composition shares (%); 30q70 for NCD and CMNN; YLL, YLD, YLL/YLD ratio. Joinpoint-regression AAPC with 95% CI.

| Metric | **1990** | **2000** | **2010** | **2023** | **AAPC_%** | **AAPC_95CI** |
|---|---|---|---|---|---|---|
| CMNN age-std DALY rate (per 100k) | 13295.9 | 8549.0 | 5870.2 | 4022.1 | -4.63 | -4.80, -4.46 |
| NCD age-std DALY rate (per 100k) | 21688.2 | 19960.4 | 19742.8 | 19282.8 | -0.37 | -0.45, -0.30 |
| Injuries age-std DALY rate | 5942.0 | 5409.9 | 4683.0 | 3981.8 | -1.13 | -1.24, -1.03 |

| | | | | | | |
|---|---|---|---|---|---|---|
| (per 100k) | | | | | | |
| CMNN share of total DALYs (%) | 32.49 | 25.2 | 19.38 | 14.74 | -3.44 | -3.54, -3.33 |
| NCD share of total DALYs (%) | 52.99 | 58.85 | 65.17 | 70.67 | 1.09 | 1.04, 1.15 |
| Injuries share of total DALYs (%) | 14.52 | 15.95 | 15.46 | 14.59 | 0.17 | 0.03, 0.30 |
| CMNN/NCD ratio | 0.613 | 0.428 | 0.297 | 0.209 | -4.41 | -4.57, -4.25 |
| 30q70 NCD probability (%) | 25.02 | 22.76 | 22.7 | 21.8 | -0.25 | -0.33, -0.16 |
| 30q70 CMNN probability (%) | 6.18 | 3.98 | 3.29 | 2.12 | -5.86 | -6.25, -5.47 |
| YLL rate (per 100k) | 30304.5 | 23903.6 | 20687.4 | 17687.7 | -1.79 | -1.87, -1.71 |
| YLD rate (per 100k) | 10621.7 | 10015.7 | 9608.7 | 9599.0 | -0.34 | -0.37, -0.30 |
| YLL/YLD ratio | 2.85 | 2.39 | 2.15 | 1.84 | -1.51 | -1.59, -1.43 |

**Table S2.** Period-specific AAPC for Vietnam: full 1990-2023 window (BIC-selected joinpoint) versus three decade sub-periods (1990-2000, 2000-2010, 2010-2023; single-segment log-linear). 95% CI and p-value for every metric × window.

| Cause | period | aapc | ci_low | ci_high | p_value | n_obs |
|---|---|---|---|---|---|---|
| DALY - CMNN | 1990-2023 | -4.628 | -4.796 | -4.461 | 0.0 | 34 |
| DALY - CMNN | 1990-2000 | -4.354 | -4.617 | -4.091 | 0.0 | 11 |
| DALY - CMNN | 2000-2010 | -3.575 | -3.778 | -3.372 | 0.0 | 11 |
| DALY - CMNN | 2010-2023 | -2.037 | -3.242 | -0.817 | 0.006844 | 14 |
| DALY - NCD | 1990-2023 | -0.375 | -0.447 | -0.303 | 0.0 | 34 |
| DALY - NCD | 1990-2000 | -0.856 | -0.887 | -0.824 | 0.0 | 11 |
| DALY - NCD | 2000-2010 | -0.095 | -0.112 | -0.079 | 1e-06 | 11 |
| DALY - NCD | 2010-2023 | -0.277 | -0.376 | -0.178 | 0.00014 | 14 |
| DALY - Injuries | 1990-2023 | -1.134 | -1.238 | -1.03 | 0.0 | 34 |
| DALY - Injuries | 1990-2000 | -0.863 | -1.124 | -0.603 | 0.000116 | 11 |
| DALY - Injuries | 2000-2010 | -1.545 | -1.653 | -1.437 | 0.0 | 11 |
| DALY - Injuries | 2010-2023 | -1.365 | -1.474 | -1.257 | 0.0 | 14 |
| YLL rate - | 1990- | -1.793 | -1.873 | -1.713 | 0.0 | 34 |

| | | | | | | |
|---|---|---|---|---|---|---|
| All causes | 2023 | | | | | |
| YLL rate - All causes | 1990-2000 | -2.372 | -2.474 | -2.27 | 0.0 | 11 |
| YLL rate - All causes | 2000-2010 | -1.408 | -1.493 | -1.324 | 0.0 | 11 |
| YLL rate - All causes | 2010-2023 | -1.088 | -1.291 | -0.884 | 0.0 | 14 |
| YLD rate - All causes | 1990-2023 | -0.335 | -0.375 | -0.296 | 0.0 | 34 |
| YLD rate - All causes | 1990-2000 | -0.583 | -0.654 | -0.512 | 0.0 | 11 |
| YLD rate - All causes | 2000-2010 | -0.414 | -0.431 | -0.397 | 0.0 | 11 |
| YLD rate - All causes | 2010-2023 | -0.029 | -0.101 | 0.042 | 0.438574 | 14 |

**Table S3.** Das Gupta decomposition of 1990-to-2023 absolute DALY change, Vietnam, NCD and CMNN separately. Population-size, age-structure (ageing), and age-specific-rate effects in absolute DALYs; residuals below $10^{-9}$.

| cause_group | pop_size | age_structure | age_rate | total_decomp | observed_change | residual | direction |
|---|---|---|---|---|---|---|---|
| CMNN | 3247842.317505668 | -1508711.0383479963 | -8184519.739394551 | -6445388.460236879 | -6445388.46023688 | -9.313225746154785e-10 | decrease |
| NCD | 6257107.694348078 | 6080731.321756319 | -1710854.58893362 | 10626984.427170772 | 10626984.427170772 | 0.0 | increase |

| | | | |
|---|---|---|---|
| | | | 48 |

**Table S4.** Southeast-Asian NCD-share ranking, delta, joinpoint AAPC, and SDI obs/exp ratio (2023). 11 countries sorted by 2023 NCD share.

| country | ncd_share_1990 | ncd_share_2000 | ncd_share_2010 | ncd_share_2023 | delta_ncd_share_pp | aapc_ncd_share | aapc_ncd_share_lo | aapc_ncd_share_hi | aapc_ncd_share_n_bkps | ncd_share_expected_2023 | obs_vs_expected_ratio_2023 |
|---|---|---|---|---|---|---|---|---|---|---|---|
| Brunei | 76.11659158671166 | 78.7945037397907 | 80.24990537780168 | 81.26307285134496 | 5.1464812646332945 | 0.22497050611989255 | 0.1699114895311116 | 0.2800597862405052 | 2 | 77.30375892592232 | 1.0512176119303165 |
| Singapore | 78.04675389967127 | 79.22172374651193 | 79.56448277227469 | 79.28415827647095 | 1.2374043767996739 | 0.09549251102414225 | 0.044099296689026346 | 0.14691212634139195 | 1 | 83.87540014403909 | 0.9452611628715498 |
| Malaysia | 67.39564380283922 | 71.98046474715956 | 72.48321560972057 | 75.04206721646014 | 7.646423413620923 | 0.8428073115164469 | 0.6787316824133693 | 1.0071503338575427 | 3 | 74.69977047907338 | 1.0045822997204878 |
| Vietnam | 52.9935718427928 | 58.8467708847 | 65.1664658887220 | 70.6673095464069 | 17.6737377036141 | 1.09438715155463 | 1.035699910657328 | 1.15310848131602 | 3 | 67.08401324738179 | 1.053415055623033 |

| | | | | | | | | | | | |
|---|---|---|---|---|---|---|---|---|---|---|---|
| | 4 | 2648 | 3 | 7 | 3 | 22 | 4 | 3 | | | |
| Thailand | 61.324618785625 75 | 61.73344976189057 | 66.49912809951391 | 68.37638266871126 | 7.05176388308550 6 | 0.45456169399360 48 | 0.350295077035922 55 | 0.55893664673050 13 | 3 | 70.55399260618 01 | 0.969135553396334 7 |
| Indonesia | 46.91674346022182 4 | 55.17706019402696 | 62.958147169 9717 | 68.036901018205 37 | 21.12015755798354 6 | 0.77388193720546 73 | 0.458552149795665 7 | 1.09020151465999 42 | 3 | 69.21878945499107 | 0.982925323512711 7 |
| Philippines | 55.08924653433594 | 62.46890859686645 | 65.138447941577 43 | 67.34309769527 3 | 12.25385116093706 | 0.98491671405835 92 | 0.802531574607923 1 | 1.16763184858503 79 | 3 | 69.65055742534133 | 0.966870907924294 8 |
| Myanmar | 44.73297886676415 | 51.99316051415097 | 59.20150636130349 | 65.38636829533303 | 20.65338942856888 | 0.62043977578445 38 | 0.525974607633861 2 | 0.71499371370848 94 | 3 | 62.1880193338101 96 | 1.051430307570255 1 |
| Cambodia | 37.28145009262366 | 40.83213426741728 | 54.99447786958785 4 | 63.82860236923356 5 | 26.54715227660990 7 | 1.82333005391530 63 | 1.575247002919000 2 | 2.07201901237437 1 | 3 | 60.53130344866595 | 1.054472623794792 6 |

| | | | | | | | | | | | |
|---|---|---|---|---|---|---|---|---|---|---|---|
| Timor-Leste | 36.12823750797305 | 44.40753124040688 | 52.00875981548525 | 59.58314066090546 | 23.45490315293241 | 0.7899734281438642 | 0.5899713990868971 | 0.9903731192248744 | 3 | 61.57877317129889 | 0.9675922008897122 |
| Lao PDR | 28.73020505168182 | 37.29389509293426 | 49.742781171372165 | 58.64911008662491 | 29.918905034943087 | 2.299488574014208 | 2.1436504644882337 | 2.455564441993796 | 3 | 62.95900323498919 | 0.9315444507233069 |

**Table S5.** Southeast-Asian YLL/YLD ratio, 2023. 11 countries sorted by YLL/YLD descending.

| **Country** | **yll_rate** | **yld_rate** | **ratio** |
|---|---|---|---|
| Lao PDR | 36318.425358330416 | 9841.03874618124 | 3.6905073026385122 |
| Myanmar | 37840.652699147126 | 10830.66253547372 | 3.4938446817272215 |
| Timor-Leste | 28566.54316668732 | 10353.73705652385 | 2.7590562722169625 |
| Philippines | 23919.56614215732 | 10228.16501241106 | 2.338597990268327 |
| Indonesia | 24038.42791717309 | 10280.86294186098 | 2.338172199465369 |
| Cambodia | 22510.4784725558 | 10565.87167412551 | 2.1304894822526688 |
| Vietnam | 17687.694041206145 | 9599.01309002062 | 1.8426575602438466 |
| Thailand | 16483.827257309484 | 10205.09248532006 | 1.6152550582977403 |
| Malaysia | 15691.152937232326 | 9996.74651545982 | 1.5696259691056678 |
| Brunei Darussalam | 13758.47215270596 | 10248.02437587526 | 1.342548734085235 |
| Singapore | 6276.737544765727 | 9426.56418993214 | 0.6658563415363442 |

**Table S6.** CMNN sub-group sensitivity analysis, Vietnam 1990-2023. Age-standardised DALY rates for full CMNN, communicable-only, and M+N+N sub-groups, joinpoint AAPC.

| Group | 1990 | 2000 | 2010 | 2023 | AAPC_% | AAPC_95 CI |
|---|---|---|---|---|---|---|
| CMNN (full, main analysis) | 13295.9 | 8549.0 | 5870.2 | 4022.1 | -4.63 | -4.80, -4.46 |
| Communicable only (sensitivity) | 8908.8 | 5611.2 | 4001.6 | 2688.2 | -5.05 | -5.24, -4.86 |
| Maternal+Neonatal+Nutritional (sensitivity) | 4387.1 | 2937.8 | 1868.6 | 1334.0 | -3.45 | -3.66, -3.25 |
| Non-communicable diseases | 21688.2 | 19960.4 | 19742.8 | 19282.8 | -0.37 | -0.45, -0.30 |
| Injuries | 5942.0 | 5409.9 | 4683.0 | 3981.8 | -1.13 | -1.24, -1.03 |
| NCD share vs full CMNN (main analysis, % of total DALYs) | 52.99 | 58.85 | 65.17 | 70.67 | | |
| NCD share vs C-only CMNN (% of total DALYs) | 59.36 | 64.43 | 69.45 | 74.3 | | |
| NCD share vs M+N+N only CMNN (% of total DALYs) | 67.74 | 70.51 | 75.08 | 78.39 | | |

**Table S7.** Premature NCD mortality (30q70, strict WHO SDG 3.4.1 definition), 11 Southeast-Asian countries, 1990-2023. Point estimates; absolute change; relative reduction; AAPC (log-linear + joinpoint with 95% CI); n joinpoints. Vietnam ranks 6th of 11 in 2023.

| Country | 30q70_199 | 30q70_200 | 30q70_201 | 30q70_202 | delta_ab | rel_reduction_pct | AAPC_loglin | AAPC_loglin_CI | AAPC_joinpoin | AAPC_joinpoint_C | n_joinpoints |
|---|---|---|---|---|---|---|---|---|---|---|---|

| | **0** | **0** | **0** | **3** | **s** | | | | **t** | **l** | |
|---|---|---|---|---|---|---|---|---|---|---|---|
| Vietnam | 22.166 | 20.196 | 20.199 | 19.504 | -2.662 | 12.01 | -0.366 | -0.42, -0.31 | -0.366 | -0.42, -0.31 | 0 |
| Thailand | 21.236 | 22.18 | 17.115 | 15.465 | -5.771 | 27.18 | -1.438 | -1.65, -1.23 | -1.438 | -1.65, -1.23 | 0 |
| Indonesia | 24.832 | 23.596 | 26.461 | 25.101 | 0.269 | -1.08 | 0.336 | 0.23, 0.45 | 0.336 | 0.23, 0.45 | 0 |
| Philippines | 21.733 | 23.762 | 24.968 | 23.179 | 1.446 | -6.66 | 0.327 | 0.22, 0.44 | 0.327 | 0.22, 0.44 | 0 |
| Malaysia | 23.625 | 23.069 | 19.517 | 18.575 | -5.05 | 21.38 | -0.863 | -0.98, -0.75 | -0.863 | -0.98, -0.75 | 0 |
| Myanmar | 35.898 | 35.258 | 32.592 | 32.394 | -3.504 | 9.76 | -0.45 | -0.52, -0.38 | -0.45 | -0.52, -0.38 | 0 |
| Cambodia | 21.067 | 19.542 | 19.325 | 19.283 | -1.783 | 8.47 | -0.363 | -0.42, -0.31 | -0.363 | -0.42, -0.31 | 0 |
| Lao PDR | 26.201 | 27.435 | 27.419 | 29.48 | 3.278 | -12.51 | 0.227 | 0.18, 0.27 | 0.227 | 0.18, 0.27 | 0 |
| Singapore | 22.457 | 16.195 | 10.634 | 7.683 | -14.775 | 65.79 | -3.6 | -3.71, -3.49 | -3.6 | -3.71, -3.49 | 0 |
| Brunei | 26.806 | 22.802 | 18.619 | 16.73 | -10. | 37.59 | -1.61 | -1.70, -1.53 | -1.61 | -1.70, -1.53 | 0 |

| | | | | | | | | | | | |
|---|---|---|---|---|---|---|---|---|---|---|---|
| Darussalam | | | | | 076 | | | | | | |
| Timor-Leste | 23.358 | 22.312 | 19.904 | 22.128 | -1.23 | 5.27 | -0.219 | -0.37, -0.07 | -0.219 | -0.37, -0.07 | 0 |

**Table S8.** Southeast-Asian NCD death rate 2023. Age-standardised deaths per 100,000 from level-1 NCD aggregate across 11 SEA countries with 95% UI, sorted ascending.

| **Country** | **ncd_death_rate_asr** | **ncd_death_rate_asr_lo** | **ncd_death_rate_asr_hi** |
|---|---|---|---|
| Myanmar | 926.887786806412 | 859.6402198128544 | 994.561771768616 |
| Lao PDR | 767.6805954997578 | 702.428050796416 | 839.0238737267475 |
| Timor-Leste | 639.6616715204299 | 580.4531353849326 | 707.3808800816366 |
| Philippines | 615.2632876782724 | 596.1866994571551 | 635.0587494920491 |
| Indonesia | 567.7801039018887 | 528.4294067673587 | 618.7111440509509 |
| Cambodia | 540.6278521896644 | 495.5591759571849 | 585.1639375977696 |
| Malaysia | 501.0951353927223 | 477.0476291548605 | 526.856410574241 |
| Vietnam | 492.2771357293784 | 450.5135126832341 | 531.3239705648899 |
| Brunei Darussalam | 491.1008240010142 | 468.5083243443163 | 513.5532754939491 |
| Thailand | 406.05308019287503 | 393.53900114258346 | 420.1797472535745 |
| Singapore | 235.99560911953583 | 226.4360712593967 | 247.02804554638595 |

**Table S9. GBD 2023 cause-hierarchy reference used in this analysis.**

| Level | Cause ID | Cause name | Used in |
|---|---|---|---|
| 1 | 294 | All causes | Denominator for composition shares |
| 1 | 295 | CMNN | §3.1, §3.2, §3.8, Figs 1, 2, S4 |
| 1 | 409 | NCD | §3.1-§3.5, all figures |
| 1 | 687 | Injuries | §3.1, Fig 1 |
| 2 | 491 | Cardiovascular diseases | SDG 3.4.1 30q70 (§3.5, Fig 4, Table S7) |
| 2 | 410 | Neoplasms | SDG 3.4.1 30q70 |
| 2 | 627 | Diabetes and kidney diseases | SDG 3.4.1 30q70 |
| 2 | 508 | Chronic respiratory diseases | SDG 3.4.1 30q70 |
| 2 | 297 | HIV/AIDS and STIs | CMNN sensitivity (§3.8, Fig S4, Table S6) |
| 2 | 322 | Respiratory infections and tuberculosis | CMNN sensitivity |
| 2 | 339 | Enteric infections | CMNN sensitivity |
| 2 | 344 | Neglected tropical diseases and malaria | CMNN sensitivity |

| | | | |
|---|---|---|---|
| 2 | 366 | Maternal and neonatal disorders | CMNN sensitivity |
| 2 | 371 | Nutritional deficiencies | CMNN sensitivity |
| 2 | 398 | Other infectious diseases | CMNN sensitivity |

**S3. GATHER reporting statement**

This study conforms to the Guidelines for Accurate and Transparent Health Estimates Reporting (GATHER), an 18-item reporting checklist (Stevens et al. Lancet 2016;388:e19-23). The full checklist is in Table S10. Key compliance: (i) input data from single public release (GBD 2023, October 2025); (ii) open code pipeline reproducible offline under 10 min; (iii) all point estimates with 95% CI or 95% UI; (iv) all formulae in §S1 with italic notation; (v) CMNN grouping sensitivity reported transparently (§S1.7).

**Table S10. GATHER reporting checklist (18 items, Stevens et al. 2016).**

| GATHER section | Checklist item | Reported in |
|---|---|---|
| Objectives and funding | 1. Define indicator(s), populations, time period(s). | Abstract; §1; §2.1 Data source; §S1.1 |
| | 2. List funding sources. | Funding statement (main) |
| Data inputs (multiple sources) | 3. Describe data identification and access. | §2.1; §S1.1 (IHME GHDx, October 2025 release) |
| | 4. Inclusion/exclusion criteria + ad hoc exclusions. | §S1.1 (11 SEA countries, level-1 causes, 1990-2023, both-sex) |
| | 5. Information on included data sources and characteristics. | §2.1; §S1.1 (GBD 2023; 204 countries; both sexes; <1-95+; 1990-2023) |
| | 6. Categories of input data with potentially important | §4.4 Limitations (verbal autopsy for low-income settings; CMNN bundling |

| | | |
|---|---|---|
| | biases. | §S1.7) |
| Data inputs (non-synthesised) | 7. Describe and give sources for other data inputs. | §S1.1 ancillary: SDI (IHME covariate), cause hierarchy |
| Data inputs (shareability) | 8. Provide all input data in an efficient extractable format. | Raw CSVs re-extractable from GHDx via §S1.1 queries (IHME Free-of-Charge Non-commercial User Agreement precludes redistribution) |
| Data analysis | 9. Conceptual overview of analysis method. | §2.2-§2.7; §S1.2-§S1.7 |
| | 10. Detailed description of all steps, with mathematical formulae. | §S1.3 joinpoint (5 eqs); §S1.4 Das Gupta (3 eqs); §S1.5 SDI LOO; §S1.6 Chiang II (2 eqs) |
| | 11. How candidate models were evaluated and final model selected. | §S1.3 BIC-penalised dynamic programming for break-points |
| | 12. Evaluation of model performance + sensitivity analysis results. | §2.3; §3.3 (Das Gupta residuals below $10^{-9}$); §3.8 CMNN sensitivity; §S1.7; Figure S4; Table S6 |
| | 13. Methods for calculating uncertainty; sources included/excluded. | §S1.3 Clegg 2009 delta-method CI; 95% UI from GBD ensemble posterior (CODEm + DisMod-MR 2.1); §4.4 SDI LOO residual uncertainty (bootstrap CI ±0.08 not in main tables) |

| | | |
|---|---|---|
| | 14. How analyses take bias into account. | §S1.5 leave-one-out quadratic SDI fit; §S1.7 CMNN sub-group sensitivity |
| Results and discussion | 15. Published estimates in efficient extractable format. | Tables S1-S10 + raw CSV outputs in repository `tables/` |
| | 16. Quantitative uncertainty measure (e.g., UI). | 95% CI on all AAPCs (Tables S1-S4, S7); 95% UI preserved in GBD sources |
| | 17. Interpret in light of existing evidence. | §4.2 (GBD 2023 profiles; Nguyen & Hoang 2022; Tran et al. 2025 stroke; GBD 2021 ASEAN CVD; Nature Comms 2026 paradox of progress) |
| | 18. Discuss limitations: modelling assumptions and data limitations. | §4.4 Limitations (GBD UI not propagated; SDI-peer n=10 imprecision; level-1 analysis; single-segment log-linear for cross-country AAPCs) |
